# Patient-specific subnetwork connectivity predicts deep brain stimulation response in obsessive-compulsive disorder

**DOI:** 10.64898/2026.10.19.26365556

**Authors:** Robert L. Seilheimer, Nikolaos Vardalakis, Wonkyung Choi, Liming Qiu, Gustavo Campos, Young-Hoon Nho, Hengji Chen, Sergey Shuvaev, Angela M. Noecker, Andrew Chang, Cameron C. McIntyre, Desmond J. Oathes, Taneeta M. Ganguly, Mario A. Cristancho, Lily A. Brown, Daniel AN Barbosa, Bijan Pesaran, Casey H. Halpern, Katherine W. Scangos

## Abstract

Deep brain stimulation (DBS) can be an effective treatment for severe obsessive-compulsive disorder (OCD), but response is variable, and objective, reliable predictors of who will respond positively are absent. In three individuals with OCD, we implanted intracranial electrodes across the cortico-striato-limbic network before DBS. Prefrontal gamma activity, most prominently in the orbitofrontal cortex (OFC), distinguished high-from low-symptom OCD states reproducibly across days. Acute stimulation of the ventral capsule/ventral striatum relieved OCD symptoms and, in one patient, significantly shifted this OFC gamma signal. Effective connectivity from therapeutic stimulation sites to these prefrontal regions defined three patient-specific OCD subnetworks; the baseline strength of the nucleus accumbens/ventral pallidum-OFC subnetwork predicted acute and long-term DBS response. Imaging-based modeling of current propagation in stimulation-activated ventral capsule to the OFC tracked acute response, suggesting that this prediction could be approximated from imaging alone. These findings outline a predictive, subnetwork-level framework for personalized DBS in OCD.

**Highlights:**

- Prefrontal gamma encodes OCD symptom state and persists across recording days
- Acute VC/VS stimulation relieves symptoms and can shift the prefrontal gamma signal
- Baseline subnetwork connectivity predicts acute and long-term DBS response
- Modeled ALIC-to-OFC tract activation approximates this prediction from imaging

**In Brief:** Scangos and colleagues mapped three patients with obsessive-compulsive disorder with intracranial electrodes before deep brain stimulation. Prefrontal gamma activity tracked symptom state, and baseline connectivity from stimulation sites to these prefrontal regions predicted acute and long-term stimulation response. Modeled activation of ALIC-to-OFC tracts was associated with response, offering a route to this prediction from imaging alone.

**Graphical Abstract:** 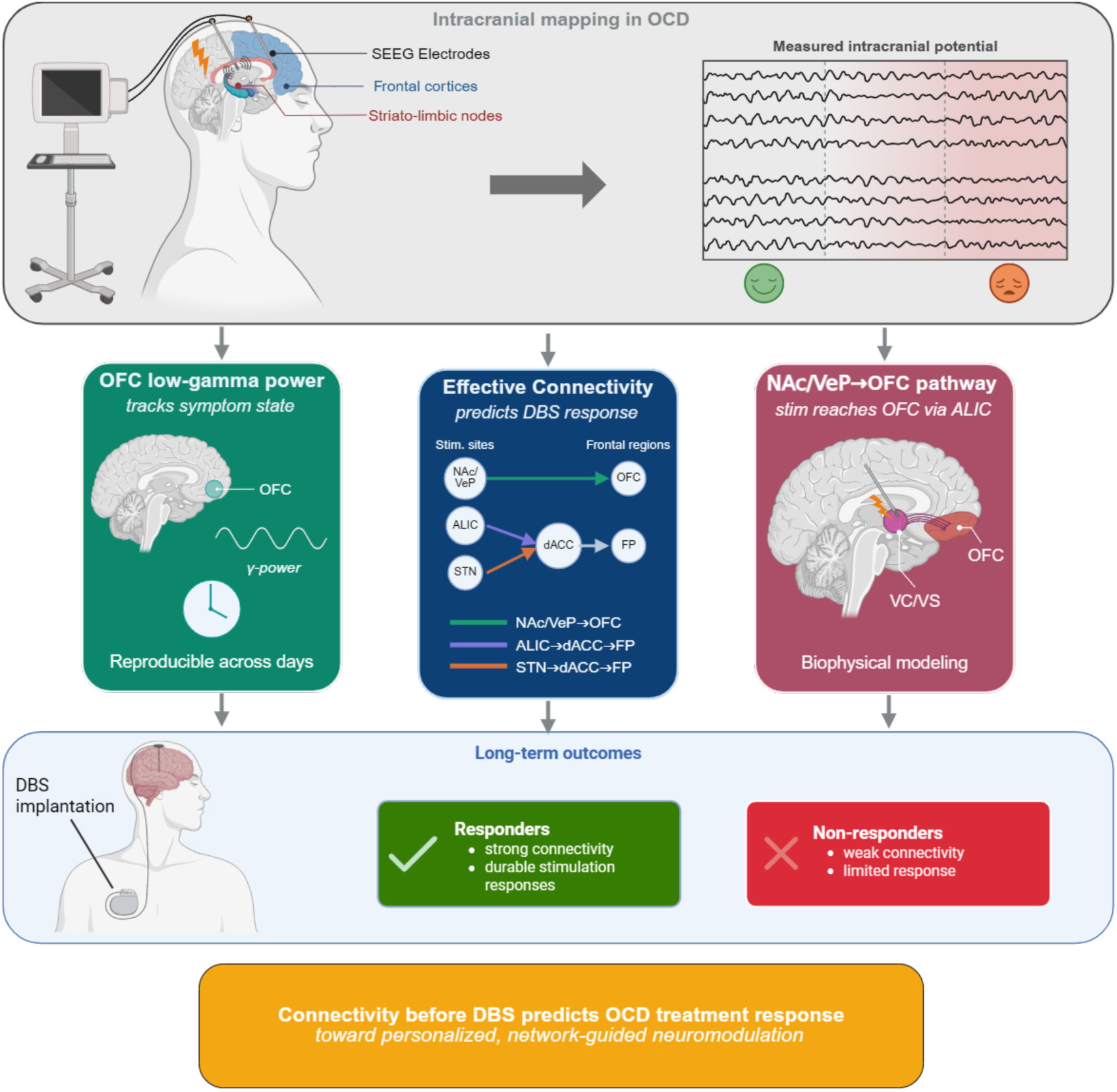

## Introduction

Obsessive-compulsive disorder (OCD) is a common, chronic, and frequently disabling psychiatric illness, affecting about 2-3% of adults worldwide^1^, including approximately 2.2 million U.S. adults with more than 200,000 new cases annually^2^. About half of patients do not reach remission despite serotonergic medications, exposure and response prevention therapy, and transcranial magnetic stimulation^3,4^. For these patients, neurosurgical intervention can be pursued^5^, and often deep brain stimulation (DBS) is considered given its reversibility and titratability. DBS of the anterior limb of the internal capsule (ALIC) and nearby nucleus accumbens (NAc) region, or of the anteromedial subthalamic nucleus (amSTN), is a developing therapy for severe, treatment-refractory OCD^6–9^. ALIC DBS received an FDA humanitarian device exemption in 2009^10^, and a recent meta-analysis reports a mean 47% reduction^11^ in the Yale-Brown Obsessive-Compulsive Scale (Y-BOCS)^12^. Yet response (≥35% Y-BOCS reduction) occurs in only about two-thirds of patients, with a moderate effect size (Hedges’ *g* 0.56)^13^, and programming is prolonged and largely trial-and-error^14^. No objective measure taken before implantation reliably identifies who will respond beyond clinical intuition.

OCD is increasingly understood as a disorder of a distributed cortico-striato-limbic (CSL) network, in which hyperactivity across the orbitofrontal cortex (OFC), anterior cingulate cortex, and limbic ventral basal ganglia is associated with obsessions and compulsions^15^. Within this network, anatomically distinct subnetworks subserve different OCD-relevant roles^16,17^. A ventral subnetwork linking the NAc and ventral pallidum (VeP) to the OFC (NAc/VeP–OFC) via ALIC appears to mediate affective processes and reward valuation^18^. Dorsal subnetworks linking the ALIC and amSTN to the dorsal anterior cingulate cortex (dACC) and frontal pole (FP) (ALIC–dACC–FP and amSTN–dACC–FP) are likely involved in cognitive control and error monitoring^19^.

Stimulation reaches these subnetworks through specific circuits. The ALIC carries prefrontal fibers in a topographic arrangement, so neighboring ALIC sites can engage different prefrontal territories including the OFC^20–22^, and the amSTN acts on the same subnetworks via hyperdirect and indirect pathways^23,24^. We therefore describe OCD at three nested levels: the CSL network as a whole; subnetworks, sets of regions within it that serve related functions and that we define in each patient by their inter-node connectivity; and circuits, the specific anatomical pathways within a subnetwork through which stimulation at a given site reaches cortex. Which subnetwork a patient’s stimulation engages, through which circuit, and how strongly nodes in this subnetwork are interconnected may therefore explain why DBS therapy helps some patients and not others.

A network view also informs where neural activity correlates of symptom states might be available as readouts. Candidate markers recorded from the DBS lead itself, including ventral-striatal theta-alpha-band power periodicity, increases in ventral-striatal delta-band power during symptom provocation, and subthalamic theta band power changes with symptoms, have been largely retrospective, group-level, or variable across patients^25–28^. We recently found that low-gamma activity in the OFC tracks moment-to-moment symptom severity and is reduced by symptom-relieving stimulation in the NAc vicinity^29^, and an independent invasive-mapping program converged on similar OFC activity as an intracranial electroencephalogram (iEEG) marker of symptom state^30^. Because these recently reported signals arise in cortex, away from where DBS is delivered, linking a stimulation site to its therapeutic effect requires understanding the connectivity between target sites to the cortical regions that carry the symptom correlate iEEG signal.

Tractography-based models of stimulation-activated pathways have been used to computationally investigate this connectivity with mixed success^31,32^. Tractography suffers from an inherent tradeoff where increasing sensitivity to detect true connections inevitably reduces specificity by generating false positives, making it difficult to accurately determine whether a specific anatomical connection exists^33^. This limitation persists even with modern algorithms, as no single method consistently identifies true pathways without also producing significant errors in reconstruction^34^. Effective connectivity (EC), measured by single-pulse electrical stimulation, offers a direct physiological alternative: a pulse delivered through one contact evokes a time-locked response elsewhere in the brain whose amplitude indexes the strength, and may index the directionality, of connectivity to other regions^35,36^. Whether single-pulse EC predicts psychiatric DBS outcome has not, to our knowledge, been tested.

Perioperative mapping with iEEG facilitated by stereo-EEG surgery, which is now routine in treatment-refractory epilepsy^37^ and recently applied to personalize and guide DBS planning for depression, chronic pain, and OCD^29,38,39^, makes it possible to measure symptom-state activity, stimulation responses and EC. In three patients mapped with stereo-EEG before DBS as part of a clinical trial of stereo-EEG-guided DBS for OCD^40^, we asked whether spectral activity distinguishing high-from low-symptom states is distributed across the CSL network and stable across days, whether acute stimulation relieves symptoms and shifts that activity, how symptom-relieving stimulation sites connect to the regions exhibiting it and whether the baseline strength of that EC predicts long-term DBS response.

We found that gamma activity, most prominently distributed across prefrontal regions, encoded symptom state and reproducibly encoded symptom state across days, and that acute stimulation at specific subcortical sites relieved symptoms. EC from these sites to the regions most prominently exhibiting iEEG symptom correlates defined three subnetworks, common across patients and identified at the individual level, and the baseline EC of the NAc/VeP–OFC subnetwork predicted both the acute and the long-term DBS response. Biophysical modeling of stimulating ALIC pathways to OFC exhibited promise toward approximating this prediction from imaging alone. These results suggest that patients and sites within these subnetworks could be selected for DBS treatment on the basis of connectivity measured before chronic implantation and potentially before any surgery at all.

## Results

### Prefrontal gamma activity encodes symptom state across the network and across days

Three adults with severe, treatment-refractory OCD (P1, P2, P3; Table S1) were implanted with stereo-EEG depth electrodes across the ALIC, NAc, VeP, amSTN, zona incerta (ZI), dACC, OFC, and FP for a 12-day inpatient mapping period^40^ (**Fig. 1a**). Throughout this period, patients rated obsessions, compulsive urges, OCD-related distress, anxiety, energy, and depression on visual analog scales (VAS; total ratings P1: 597; P2: 755; P3: 724) while iEEG activity was recorded continuously, including during provocation with tailored OCD-relevant exposures. We clustered each patient’s ratings into high- and low-severity symptom states (**Fig. 1b; Fig. S1**),^38^ which served as labels for all subsequent analyses. Because P3 reported only OCD distress ratings for much of the iEEG phase, these symptom states were instead defined by thresholding the distress ratings alone.

**Figure 1.**
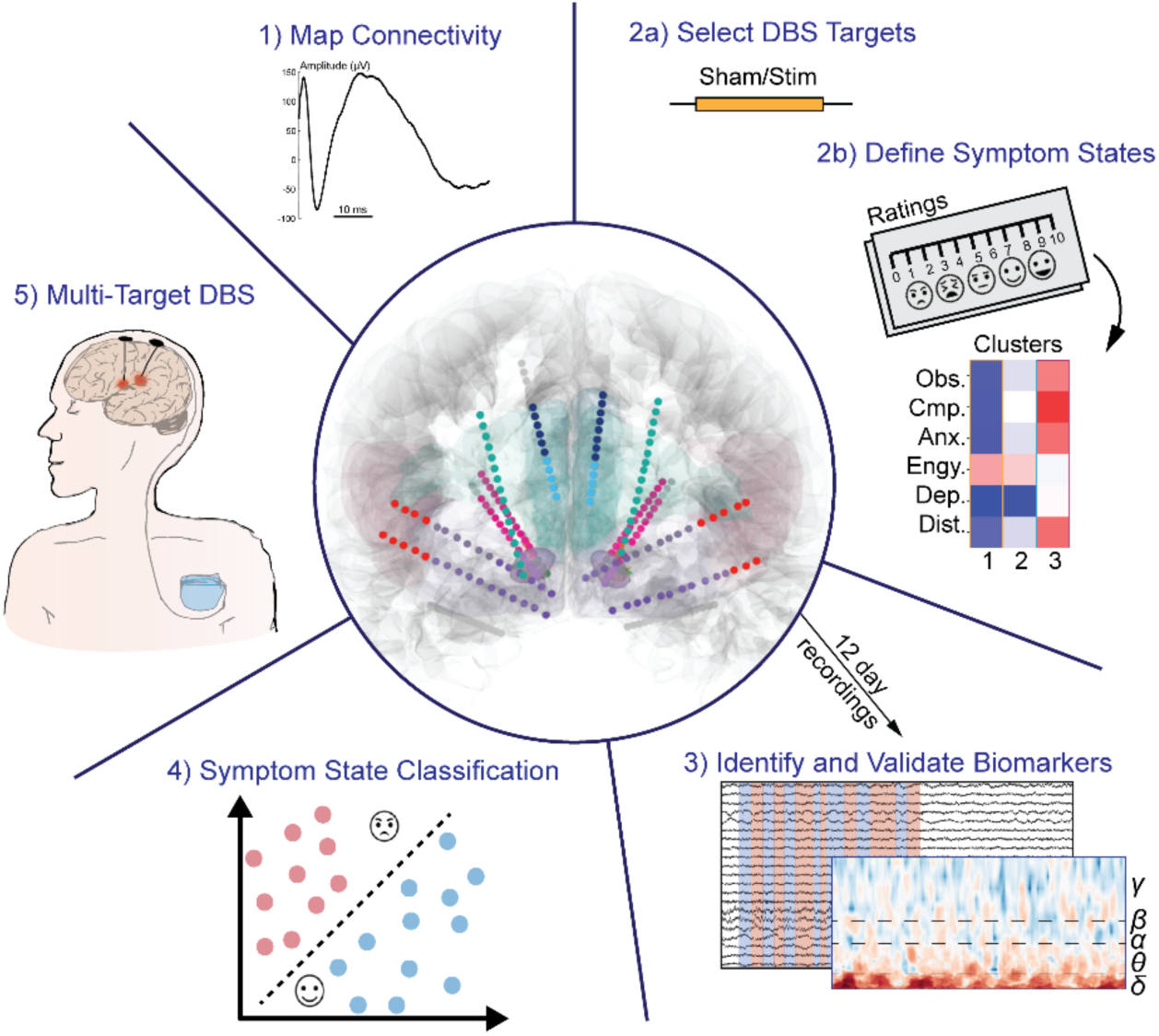
Study design and symptom states. Overview of study design, methods and procedures, and electrode locations; depth electrode contacts colored by region (see **Figure Key**). Symptom ratings clustered into high- and low-severity states for each patient. (Obs: obsessions; Cmp: compulsive urges; Anx: anxiety; Engy: energy; Dep: depression; Dist: distress).

To identify iEEG markers of these states which encoded symptom state across the implanted cortical and subcortical areas of the brain for each patient, we extracted power in six canonical frequency bands at all gray-matter contacts from stimulation-free, minute-long epochs recorded during symptom provocation (total epochs P1: 105; P2: 413; P3: 235) and trained patient-specific classifiers to distinguish high-from low-severity states. Classifiers were evaluated over 200 repetitions of grouped, stratified two-fold cross-validation with features selected at each iteration by a univariate F-test on the training half only (STAR Methods). Symptom states were reliably distinguished by these classifiers (mean AUC P1: 0.75, 2.5–97.5th percentile 0.71–0.78; P2: 0.74, 0.68–0.78; P3: 0.63, 0.54–0.71; permuted-label null 0.50, permutation p = 0.005; **Fig. 2a**).

**Figure 2.**
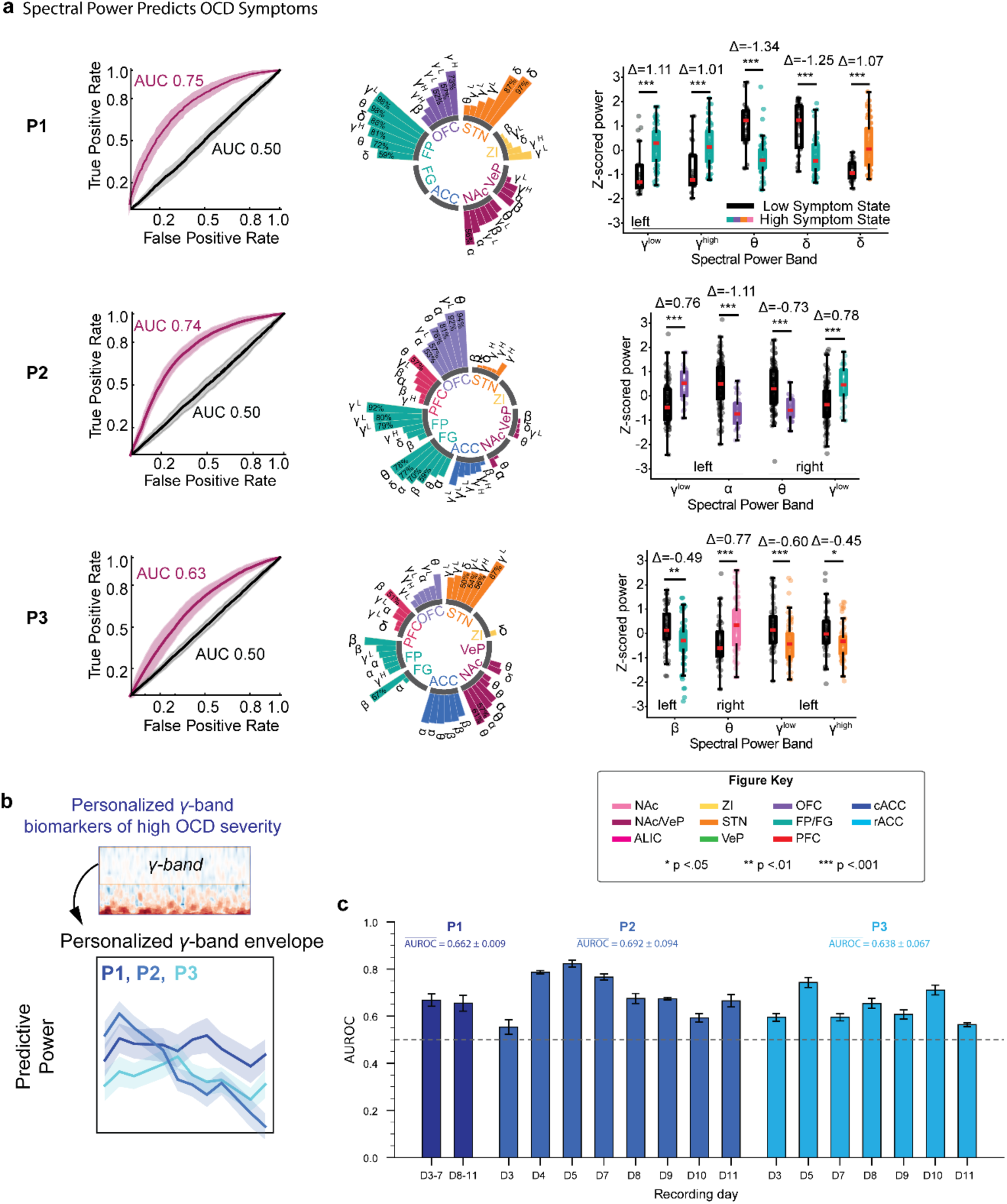
Prefrontal gamma activity encodes symptom state across days. **a**, Left, ROC curves (mean ± s.d.) for each patient’s classifier distinguishing high-vs. low-severity states (magenta) and with shuffled labels (black). Middle, top spectral-spatial biomarker features (bar height = % of runs selected, colored by region; see **Figure Key**). Right, z-scored power by region and band in high-vs. low-severity states (colored by region). Each row corresponds to one patient. **b**, Learned frequency contributions to high-symptom states from the factorized gamma-band decoder, per patient. Positive values indicate higher gamma power in high-symptom states; error bars, variability across bootstrap runs (contribution of each 10-Hz bin, 30-150 Hz). **c**, Per-day classification performance (AUROC) for each patient, with bars ordered by increasing AUROC and each patient’s mean across days in parentheses (P1 0.66, P2 0.69, P3 0.64). Error bars, standard deviation across random-seed runs; dashed line, chance (0.5).

The most informative spectral features formed distributed spectral-spatial biomarker sets spanning multiple CSL regions and were dominated by gamma-band power in the OFC and FP (**Fig. 2a**). In P1 and P2, prefrontal gamma power (low and high gamma) was higher during high-severity states (P1: p < 0.0001; P2: p = 0.0007); in P3 it did not significantly differ between states. Subthalamic activity also varied with symptom state, but in a patient-specific way (increased delta in P1, p < 0.0001; reduced gamma in P3, p = 0.0072).

Because this analysis averaged power within broad canonical bands and highlighted only the strongest features, we next resolved the gamma range more finely. An independent decoder with different architecture applied to the same stimulation-free provocation recordings used gamma-band power in 10-Hz bins (30–150 Hz) and learned, for each patient, a frequency envelope and a signed contribution of each region to high-symptom states (**Fig. 2b**; STAR Methods). The OFC, rostral ACC, and ventrolateral PFC iEEG contributed to high-symptom states in all three patients, with patient-specific contributions from the frontal pole (P2, P3), zona incerta (P1, P2), and ALIC (P3). Contributions were directionally heterogeneous: gamma power increased in some regions and decreased in others during high-symptom states, with corresponding regions in opposite hemispheres contributing in opposing directions (e.g., left versus right anterior medial OFC in P1). The frequency envelopes also differed, spanning the gamma range broadly in P1 and P3, and peaking in the low-gamma range near 40 Hz in P2. At this finer resolution, OFC gamma also contributed to symptom-state decoding in P3, a contribution that the band-averaged analysis did not reveal. Evaluated with iteratively stratified folds (see STAR methods), symptom states were decoded from the gamma power alone, with consistent performance across recording days (mean per-day AUROC P1: 0.66; P2: 0.69; P3: 0.64; **Fig. 2c**).

Symptom state was therefore encoded by prefrontal gamma activity that persisted across days. The OFC contributed in every patient. As these regions tracked symptom state, we then asked whether stimulation of the subcortical nodes of the network could move them.

### Acute stimulation relieves symptoms and can shift prefrontal gamma

During iEEG mapping of the CSL network, we delivered minute-long, double-blinded, sham-controlled stimulation to regions of the CSL network that have been implicated as potential stimulation or ablation targets for the treatment of OCD in prior studies, including the ALIC, NAc, VeP, amSTN, and dACC (both rostral (rACC) and caudal (cACC) subregions), and scored the immediate change on the same VAS scales, yielding a multi-dimensional response profile for each patient (**Fig. 3a**; **Fig. S2**). Each patient had a sham condition against which all targets were compared. P1 and P2 exhibited stimulated site-specific and symptom-specific improvements. In P1, right NAc/VeP^28^ stimulation reduced obsessions and compulsive urges relative to sham (ΔObs = −1.67 and ΔCmp = −1.67 vs. 0.11 and 0; p = 0.036, one-sided Wilcoxon rank-sum test), and left NAc/VeP stimulation reduced compulsive urges (ΔCmp = −1.56 vs. 0; p = 0.036). In P2, the largest sham-separated reductions came from the right STN (ΔCmp = −2.71 vs. −0.22; p = 0.038) and left cACC (ΔCmp = −2.3 vs. −0.22; p = 0.004), and left NAc/VeP (ΔDis = −0.96 vs. 0.22; p = 0.005). Stimulation sites differed in the symptoms they affected: NAc/VeP stimulation also improved affect (depression in P1, energy in P2), amSTN stimulation selectively reduced OCD-related distress. No site produced sham-separated improvement in P3.

**Figure 3.**
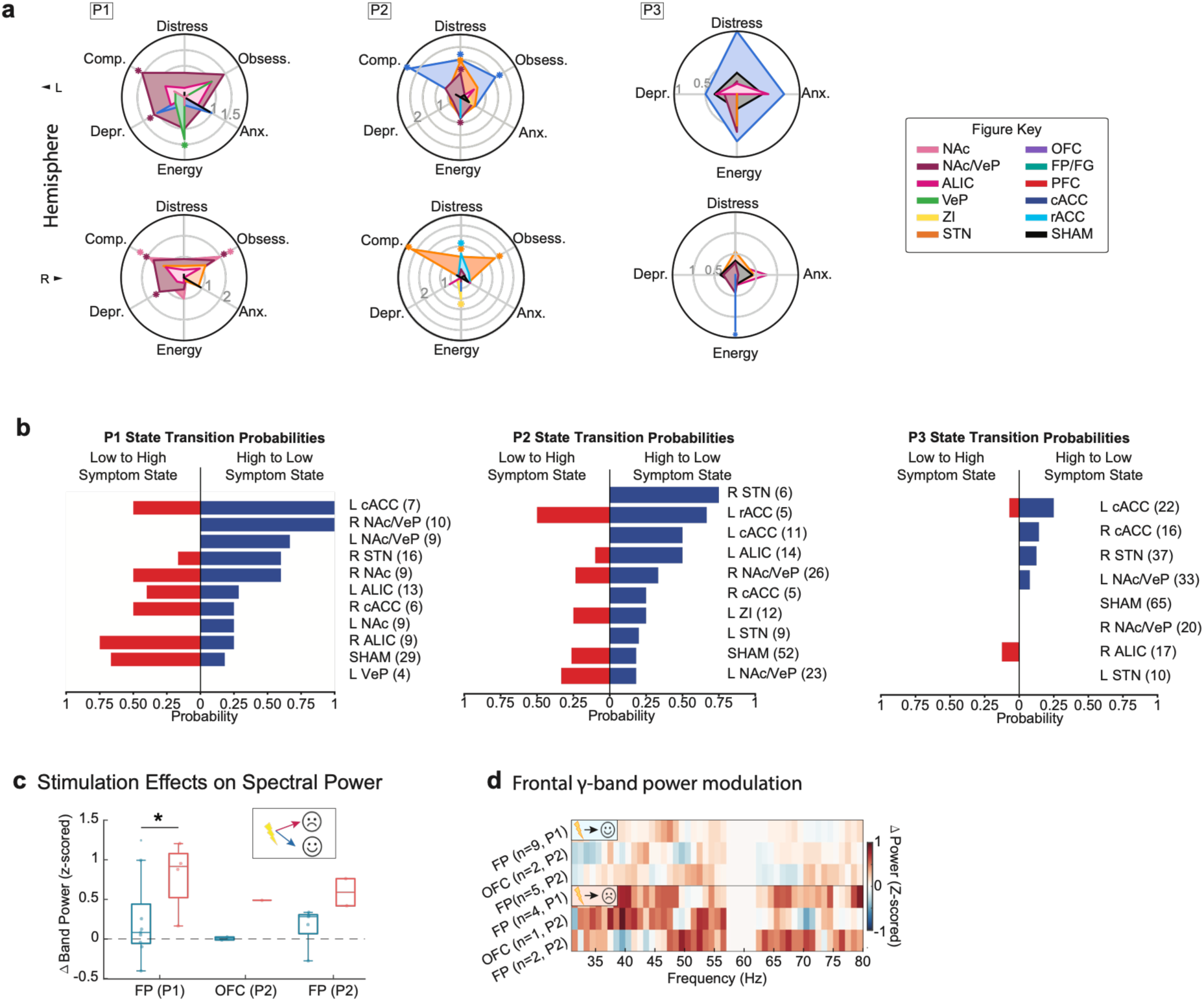
Prefrontal gamma activity tracks the behavioral effects of stimulation. **a**, Radar plots of patient- and site-specific stimulation-induced symptom improvements compared with sham; responses colored by stimulation site (see **Figure Key**). **b**, Probability of transitioning from the high-to the low-severity state (blue) and from the low-to the high-severity state (red) for each stimulation site, with the number of stimulation trials per site in parentheses. **c**, Prefrontal low-gamma power increases after stimulation trials that worsened symptom states (red) but not those that improved (blue) symptom states, shown for P1 and P2 at prefrontal electrode contacts selected in more than 90% of classifier runs. P3 had no such contact; its result is reported in the STAR Methods. **d**, Heatmap of frontal gamma power for improving vs. worsening stimulation

To compare stimulation sites on a common scale, we computed for each site the probability that stimulation moved the patient from the high-to the low-severity state (**Fig. 1b**; STAR Methods). Transition probabilities for active stimulation of select sites were generally higher than for sham in P1 and P2, though varied between patients (P1 left cACC and right NAc/VeP, 100% state improvement vs. 18% state improvement during sham; P2 right amSTN, 75% vs. 18%). In P3, the best site reached only 25% (left cACC; sham 0%; **Fig. 3b**). This transition probability served as the acute response measure in subsequent analyses; for the STN, whose effect was specific to distress, the change in distress was used instead (**Fig. 4d**).

**Figure 4.**
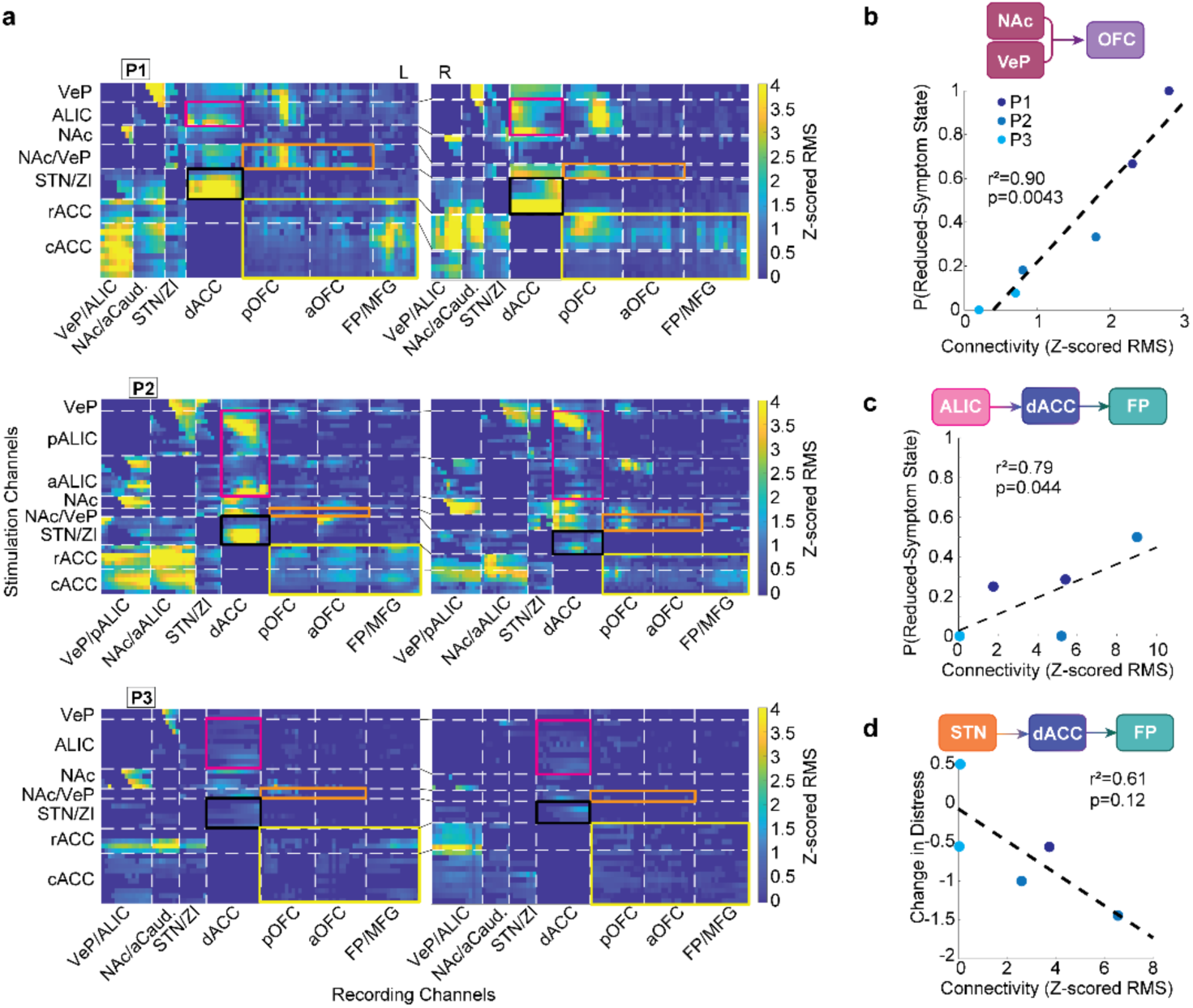
Effective connectivity predicts acute stimulation response. **a**, Heatmaps of effective connectivity from stimulation sites (vertical axis) to recording sites (horizontal axis) for each participant; highlighted connections: ALIC–dACC(maroon), NAc/VeP–OFC (orange), STN–dACC (black), dACC–OFC/FP (yellow). **b**, Effective connectivity from NAc/VeP to OFC correlates with the probability of leaving the high-severity state with NAc/VeP stimulation. **c**, Effective connectivity from ALIC to FP via dACC correlates with the probability of leaving the high-severity state with ALIC stimulation. **d**, Effective connectivity from STN to FP via dACC correlates with decrease in distress with STN stimulation (all weighted Pearson correlation).

We then asked whether the symptom effects of stimulation across sites were mirrored in the prefrontal gamma iEEG signal, as we previously observed for stimulation of the NAc/VeP region^29^. We combined stimulation targets in the pericapsular regions for this analysis. Power was measured at the prefrontal electrode contacts most frequently selected by the classifiers (STAR Methods). Prefrontal gamma power increased after stimulation trials that worsened symptom state, but not after trials that improved it. This difference was significant in P1, and in the same direction but not significant in P2 (P1: FP; p = 0.0405, two-sided permutation test; P2: FP, p = 0.1377; P3: OFC, p = 0.6953, **Fig. 3c,d** and **Fig. S3**). Because stimulating these subcortical sites changed prefrontal activity, we next examined how they are connected to the prefrontal regions carrying the biomarker.

### Effective connectivity from therapeutic sites to the biomarker regions predicts acute response

To map how the stimulation sites connect to these prefrontal regions, we measured baseline effective connectivity (EC) with single-pulse evoked potentials (SPEPs) during iEEG mapping, before chronic DBS implantation. Single pulses were delivered to each electrode contact pair, and the evoked response at every other contact was quantified as its root-mean-square amplitude (STAR Methods). This gave a direct, high-resolution measure of connectivity strength, including across the densely packed, crossing white-matter pathways of the ALIC that are difficult to resolve with diffusion tractography in individual patients (**Fig. 4a**). Starting from the stimulation sites that produced clinical improvement, we identified the prefrontal regions to which each was most strongly connected, yielding three data-driven subnetworks: NAc/VeP–OFC, ALIC–dACC–FP, and STN–dACC–FP. For the two subnetworks relayed through the dACC, strength was computed as the product of their constituent connections (for example, ALIC→dACC × dACC→FP). The prefrontal nodes of these subnetworks converged on the OFC and FP regions that exhibited the gamma biomarker (**Figs. 2a,b** and **4a**).

Baseline EC within these subnetworks scaled with acute clinical response. Connectivity within the NAc/VeP–OFC and ALIC–dACC–FP subnetworks correlated with the probability of leaving the high-severity state (NAc/VeP–OFC: r² = 0.90, p = 0.0043; ALIC–dACC–FP: r² = 0.79, p = 0.044; weighted Pearson correlation; **Fig. 4b,c**; **Fig. S4**). Connectivity within the STN–dACC–FP subnetwork showed a consistent but non-significant trend with change in distress (r² = 0.61, p = 0.12; **Fig. 4d**). Because these correlations were computed across six implanted hemispheres (two per patient), we tested them with exact permutation testing over all 720 label permutations. The NAc/VeP–OFC association survived (p = 0.0014, the smallest attainable), whereas the ALIC–dACC–FP association did not (p = 0.32). Both associations were stable, assessed through cross-validation when each hemisphere was left out (mean R² = 0.90, range 0.87–0.97, and 0.73, range 0.49–0.91, respectively) and when each patient was left out (mean R² = 0.91, range 0.84–0.97, and 0.72, range 0.49–0.90). Stimulation of other sites, notably the dACC, also evoked large responses in NAc/VeP and ALIC contacts (**Fig. 4a**); these responses did not scale with acute ratings and were not analyzed further.

Baseline EC from therapeutic stimulation sites to the regions with prominent gamma activity therefore correlated with acute stimulation response, robustly so for the NAc/VeP–OFC subnetwork. We next tested whether the baseline strength of these connections also predicted long-term improvement from chronic DBS.

### Baseline subnetwork connectivity predicts long-term DBS response

Following stereo-EEG electrode explantation, each patient received a four-lead DBS system targeting symptom-relieving sites within the identified subnetworks in the hemisphere with the strongest EC within the NAc/VeP–OFC and ALIC–dACC–FP subnetworks (**Fig. 5a**). Over one year of follow-up (60 weeks for P1; last common timepoint, 32 weeks), P1 and P2, the two patients with robust acute responses, improved rapidly and durably (P1: 43% reduction in Y-BOCS-II at 14 weeks; P2: 64% at 2 weeks, 55% at 32 weeks; **Fig. 5b**), and were considered DBS responders. In these patients, stimulating the NAc/VeP and ipsilateral cACC nodes was sufficient for a robust response; in P1, adding the contralateral ALIC and STN nodes produced further benefit (55% reduction at 60 weeks). P3 showed only transient benefit (24% reduction at week 6; **Fig. 5b**) and was classified as a non-responder to DBS treatment.

**Figure 5.**
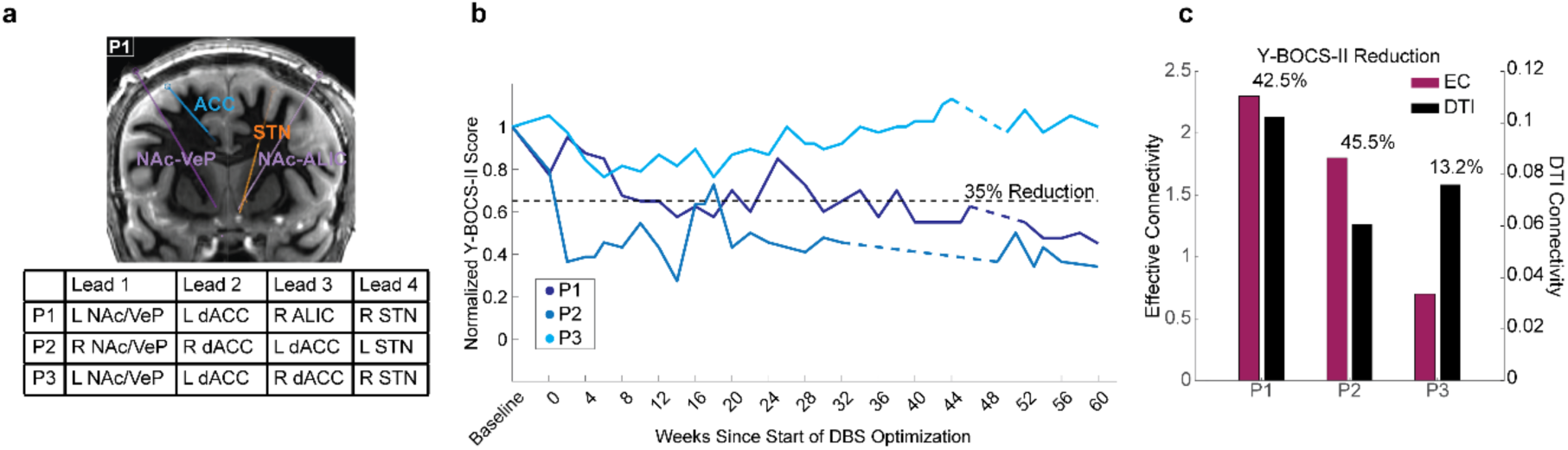
Baseline connectivity predicts long-term DBS response. **a**, CT image showing DBS lead placement for P1 and table listing lead placement for each patient. **b**, Normalized Y-BOCS-II score over the course of DBS programming, colored by patient. **c**, Strength of left NAc/VeP–posterolateral OFC (plOFC) effective connectivity (maroon) corresponds with percent reduction in Y-BOCS-II at the end of optimization of the first implanted hemisphere, whereas DTI-based connectivity (black) does not.

Across patients, the magnitude of long-term Y-BOCS-II reduction or lack thereof, measured after optimization of the NAc/VeP and ipsilateral cACC electrodes (which always occurred before optimizing other electrodes), scaled with baseline EC within the NAc/VeP–OFC subnetwork ipsilateral to the implanted NAc/VeP electrode, specifically its connection to the posterolateral OFC (plOFC; **Fig. 5c**), the same connection that predicted acute response during the iEEG mapping phase.

Long-term benefit followed the same rank order across three measures obtained during mapping (P1 ≈ P2 > P3). Responders had the strongest NAc/VeP–OFC connectivity and the most accurate symptom-state decoding (mean AUC 0.75 and 0.74 vs. 0.63), and the clearest stimulation-induced shifts in prefrontal gamma, which moved with symptom change in the same direction in both patients (significantly so in P1). In contrast, the non-responder had the weakest connectivity, the weakest decoding performance, and the smallest stimulation-induced shifts. Variability in DBS response thus appears to reflect patient-specific differences in subnetwork connectivity and in how reliably prefrontal gamma tracks symptom state and responds to stimulation, all measurable before chronic stimulation began. Since EC mapping necessitates a separate invasive procedure, we finally tested whether the same prediction could be approximated from imaging data alone.

### Imaging-derived tractographic activation approximates the connectivity predictor

We next tested whether an entirely non-invasive, imaging-based method could confirm our EC findings using patient-specific tractography. This broadly recapitulated known pathways^41^, but structural connectivity between subnetwork nodes (normalized diffusion tensor imaging waytotal) did not capture key subnetwork sites or correlate with acute response (**Fig. 5c; Fig. S5**). We therefore turned to a high-resolution, stylized connectome of the human internal capsule^42^ warped non-linearly into each patient’s space and coupled with biophysical models of axonal activation to simulate the tracts activated by each tested stimulation setting^43^. For each NAc/VeP contact, we computed the proportion of activated ALIC tracts that travel to the OFC. These tract simulations need not connect only the NAc/VeP region and the OFC; they also likely carry other frontostriatal tracts.

Across the six implanted hemispheres, the proportion of activated ALIC-to-OFC simulations correlated with the acute clinical response (r² = 0.71, p = 0.03, weighted Pearson correlation; **Fig. 6a**). The association held under permutation testing (p = 0.007) and was stable during cross-validation testing when each hemisphere was left out (mean R² = 0.74). The modeling also revealed that neighboring stimulation sites within the ALIC and the surrounding subcortical region activate different populations (**Fig. 6b-d**), underscoring the importance of circuit-based measures.

**Figure 6.**
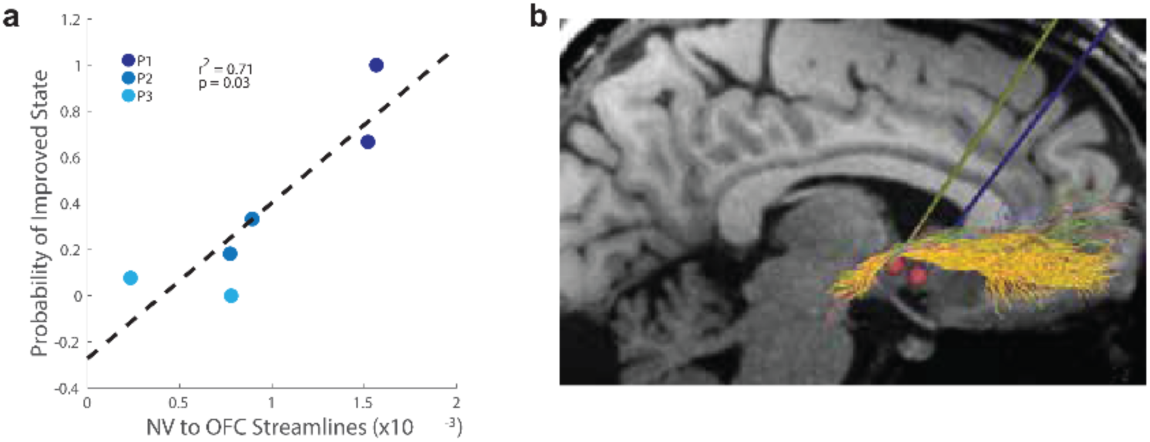
Imaging-derived tract activation tracks acute response. **a**, Proportion of activated ALIC-to-OFC tracts (biophysically modeled) engaged by NAc/VeP stimulation correlates with acute response. **b–d**, Sagittal MRI from P1 with overlaid ALIC-to-OFC tracts showing differences in tract engagement with stimulation of NAc/VeP (**b**), ventral capsule (**c**), and ventral pallidum (**d**). Yellow, OFC tracts; green, ventrolateral PFC; orange, ventromedial PFC.

Imaging-based modeling of stimulated tracts thus recovered the relationship with acute response that generic tractography missed. This suggests that pre-surgical imaging alone could predict whether a patient would respond to DBS ahead of surgery, helping with patient selection.

## Discussion

Using perioperative iEEG-based mapping in three patients with treatment-refractory OCD, we found that distributed prefrontal gamma activity encoded symptom state across the CSL network and across days, that acute stimulation at specific subcortical sites relieved symptoms and, when it worsened them, raised this activity (significantly in one patient), and that EC from those sites to the biomarker regions defined patient-specific subnetworks, of which the NAc/VeP–OFC subnetwork’s baseline strength predicted acute and long-term DBS response. Biophysical modeling of stimulation-activated ALIC-to-OFC tracts predicted the same acute response, suggesting an imaging-only route to this never-before possible outcome predictor for patients considering this invasive procedure.

The spectral findings extend prior region-level work. Our earlier study identified OFC low-gamma activity as a marker of symptom state^29^; here, symptom state was encoded by a distributed gamma signature that was stable across days and included OFC activity in each patient. However, the direction of each region’s contribution and the frequency range involved were patient-specific, arguing for individually calibrated biomarkers. Symptom state was decoded more accurately in the two responders than in the non-responder, suggesting that the functional organization of the network itself shapes therapeutic response. Subthalamic activity also tracked symptom state, but in a patient-specific way, and thus not pursued further.

Acute stimulation provided a within-session test of this relationship. The symptom effects of stimulation were mirrored in frontal gamma, which rose after trials that worsened symptoms: significantly in P1, in the same direction in P2, and close to zero in P3. In line with our earlier findings, in which NAc-vicinity stimulation reduced OFC low-gamma in all three patients ^29^, this suggests that subcortical stimulation can move the cortical signal that encodes symptom state, but in the present data the link rests on a single patient and remains associative.

The predictive value of baseline subnetwork connectivity supports a personalized, subnetwork-guided framework for DBS candidacy, target selection, and programming. Prior connectomic studies used normative structural connectivity to explain response variability retrospectively^44^, and DBS-evoked responses recorded at the stimulation site (evoked resonant neural activity and local evoked potentials) or at the cortex have helped localize optimal contacts in movement-disorder and psychiatric DBS^45–47^. Single-pulse EC instead measures how a stimulation site connects to the distant regions carrying the symptom signal. In our data, connectivity and activity measures served different purposes: connectivity assessed the capacity to respond to stimulation, whereas spectral activity tracked moment-to-moment symptom state; in our patients, how reliably it did so also appeared to track response.

These connectivity and biomarker findings share an anatomical substrate. The proportion of ALIC-to-OFC tracts activated by NAc/VeP stimulation tracked acute response. This yields a circuit-level account in which ventral ALIC stimulation relieves symptoms at least in part through the ALIC–OFC circuit that reaches the OFC regions carrying the gamma biomarker, building on prior intracranial work linking OFC activity to obsessive-compulsive behavior^29,30^. Imaging studies have long sought a connectivity-based stimulation target within the ALIC^48^. Activation of pathways to medial and lateral prefrontal cortex has predicted improvement in some cohorts^31,49,50^ but not others^32^, in part because tractography struggles to resolve the closely packed, crossing pathways of the ALIC in individual patients^51^. Consistent with this, generic waytotal connectivity did not relate to response in our patients, whereas activation-weighted modeling on a high-resolution internal capsule connectome did, suggesting that the specific fibers engaged by stimulation, rather than overall structural connectivity, carry symptom state specific signals. Because this measure is derived from imaging data augmented with the stimulation configuration rather than invasive recordings, it could in principle be computed prospectively for a planned DBS target. Whether it can stand in for SEEG-derived connectivity when selecting patients and targets is a question for future prospective studies.

These findings converge with independent invasive-mapping programs that localize OCD-related activity to the OFC and show that ventral ALIC or nearby NAc/VeP stimulation modulates it ^29,30,52^. That work framed therapeutic stimulation as a reversible “informational lesion” that transiently suppresses OFC activity during intervention, whereas here we identify a high-frequency signal in the gamma range that tracks symptom state, and additionally reveal that it is stable across days. Additionally, this tracked signal was shown to be modulated upward with interventions that worsened symptoms. Our central contribution is predictive: baseline connectivity forecasts which patients respond, even as both accounts converge on the same ALIC–OFC circuit.

The single non-responder is instructive. P3 had the weakest baseline subnetwork connectivity, the least accurate symptom-state decoding, the smallest stimulation-induced shift in frontal gamma, and only transient clinical benefit, consistent with a framework in which subnetwork connectivity and the reliability of the frontal gamma signal together govern responsiveness to stimulation. Additionally, the STN–dACC–FP subnetwork showed a concordant (though non-significant) relationship to acute response, supporting consideration of the amSTN as a personalized alternative target in patients whose connectivity favors that subnetwork^6,53^.

By integrating a patient’s symptom state estimates, information about where to stimulate, and who is likely to respond into a single mapping procedure, these findings point toward a personalized DBS pipeline to improve clinical outcomes in treatment-refractory OCD patients. Pairing connectivity-guided targeting with a stable biomarker could shorten the trial-and-error of conventional programming and, in principle, allow multi-day mapping to be condensed into intraoperative single-pulse-guided targeting or even replaced by imaging-based pathway modeling. Additionally, a personalized, stable biomarker signal that tracks symptoms in real time could further enable adaptive DBS approaches. These possibilities remain to be tested independently in future studies.

This study has limitations, largely related to the small sample, which is typical of early human invasive studies. In particular, the long-term association rests on three patients and should be read as a rank-order correspondence. Replication in larger cohorts is required to exclude overfitting and to establish predictive utility. Specifically, a simulation of imaging-only target selection in a new cohort would test whether pathway modeling alone reproduces the connectivity-based prediction. Intracranial coverage of the CSL network was extensive, but variability in electrode placement is inherent to the surgical procedure and not all relevant nodes were sampled, so subnetwork estimates may be shaped by sampling. Subnetworks were defined from the same mapping data used to measure acute response, which may inflate the acute associations; long-term DBS outcome, measured independently over the following months, is not subject to this selection. The acute analyses also treat the two hemispheres of each patient as independent observations, a simplification that could yet hold under specific circumstances since in one patient biomarker changes were observed in opposite directions per hemisphere; the associations were stable when each patient was left out, but this does not substitute for validation in an independent cohort. The pathway-activation analysis depends on biophysical modeling assumptions, including the volume of tissue activated, and assumes homogeneous tissue properties. Acute (VAS-based state transitions) and chronic (Y-BOCS-II) outcomes reflect different measures and timescales; because changes in momentary distress do not necessarily predict Y-BOCS change, we used the probability of leaving the high-severity state (defined across all six scales in P1 and P2, and by distress in P3) as the acute measure, except for the STN, whose effect was distress-specific. Finally, the relationship between stimulation and the gamma biomarker is associative; causal testing and longitudinal validation with chronic recordings will be needed.

In summary, here we found that prefrontal gamma activity encoded OCD symptom state and persisted across days; acute subcortical stimulation relieved symptoms and, when it worsened them, raised this signal, significantly in one patient; effective connectivity from therapeutic stimulation sites to the biomarker regions defined patient-specific subnetworks; and the baseline strength of the NAc/VeP–OFC subnetwork predicted acute and long-term DBS response. These findings outline a predictive, subnetwork-level framework for selecting patients and targets for DBS in OCD, and imaging-based modeling of the same tracts offers a possible route to applying it without a separate mapping surgery.

## STAR★METHODS

### KEY RESOURCES TABLE

| REAGENT or<br>RESOURCE | SOURCE | IDENTIFIER |
| --- | --- | --- |
| **Deposited data** |  |  |
| Raw and processed data, associated code | This paper | https://github.com/Halpern-Scangos-Lab/Personalized-Network-Biomarkers-of-DBS-Response-in-OCD |
| **Software and algorithms** |  |  |
| MATLAB Version: 24.2.0.3212159 (R2024b) Update 9 | The MathWorks Inc. | https://www.mathworks.com/ |
| FieldTrip (v20240620) | Oostenveld et al. 2011 | doi:10.1155/2011/156869 |
| Python versions 3.11.11 and 3.12.13 | Python Software Foundation | https://www.python.org |
| FSL version 6.0.5.1 | FMRIB Software Library | https://fsl.fmrib.ox.ac.uk/ |
| QSIPrep 0.20.1 | Cieslak et al. 2021 | doi:10.1038/s41592-021-01185-5 |
| ANTs version 2.1.0 | Tustison et al. 2021 | doi:10.1038/s41598-021-87564-6 |
| RAVE version 1.0.6 | Magnotti et al. 2020 | doi:10.1016/j.neuroimage.2020.117341 |
| BCI2000 | Schalk et al. 2004 | doi:10.1109/TBME.2004.827072 |
| **Other** |  |  |
| EEG System/Stimulator | g.tec Medical Engineering | https://www.gtec.at/ |
| sEEG electrodes | Ad-Tech | https://adtechmedical.com/ |

### RESOURCE AVAILABILITY

#### Lead contact

Further information and requests for resources should be directed to and will be fulfilled by the lead contact, Katherine W. Scangos.

#### Materials availability

This study did not generate new unique reagents.

#### Data and code availability

De-identified data and analysis code are available on GitHub (see KRT above).

### EXPERIMENTAL MODEL AND STUDY PARTICIPANT DETAILS

#### Patients and sEEG monitoring stage

We report on three patients with chronic (≥5 years since diagnosis), severe (Y-BOCS ≥28) obsessive-compulsive disorder (OCD) refractory to medications and exposure and response prevention therapy who were enrolled in a clinical trial of sEEG-guided DBS for OCD (NCT05623306). The study was approved by the US Food and Drug Administration under an investigational device exemption (IDE# G220185). All participants provided informed consent after review of the protocol with the study’s Principal Investigator. In the first stage of the trial, patients underwent 12 days of inpatient clinical monitoring during which sEEG electrodes (Ad-Tech Medical, Oak Creek, WI) were surgically implanted for intracranial stimulation and recording. Electrodes sampled the cortico-striato-limbic network, including the anterior limb of the internal capsule (ALIC), nucleus accumbens (NAc), subthalamic nucleus (STN), ventral pallidum (VeP), dorsal anterior cingulate cortex (dACC), orbitofrontal cortex (OFC), and frontal pole/frontal gyrus (FP/FG). Surgical targeting and trajectory planning have been previously described^40^. Electrode trajectories traversing the ALIC were planned to intersect streamlines projecting to prefrontal cortical regions based on probabilistic tractography and/or a previously described OCD network^54^. Postoperatively, high-resolution computed tomography (CT) was acquired and co-registered to preoperative MRI for electrode localization^55^. At the end of this stage, the sEEG electrodes were explanted.

### METHOD DETAILS

#### Symptom state identification

Symptom ratings were collected throughout the sEEG phase using custom software developed in Unity (Unity Technologies, San Francisco, CA). Patients rated current symptom severity on integer visual analog scales (0-10) for obsessions, compulsions, OCD-related distress, energy, anxiety, and depression. All scales except energy were anchored with 0 indicating absence of symptoms and 10 indicating the maximal severity. Energy was anchored with 0 indicating no energy and 10 indicating highest energy; values were inverted for analysis so that decreases correspond to clinical improvement, in line with other scales.

#### Biomarker discovery

Intracranial EEG recordings were obtained at a sampling rate of 1200 Hz using a g.HIamp amplifier (g.tec medical engineering). Preprocessing and feature extraction were performed in MATLAB using the FieldTrip toolbox^56^ (v20240620) and custom scripts. Minute-long recording segments immediately preceding symptom ratings were extracted (P1:105; P2:413; P3:235). Signals were band-pass filtered (1-150 Hz, eighth-order Butterworth), notch filtered at 60 Hz (±2 Hz) and harmonics, and downsampled to 500 Hz. Signals were re-referenced per shaft using a Laplacian montage.

Data was segmented into non-overlapping 2 second epochs. Artifact-contaminated channels and epochs were identified semi-automatically. Contacts with low signal-to-noise ratio (SNR) were detected using NoiseTools MATLAB library^57^, and epochs containing >20% low-SNR contacts were excluded. Contacts marked as low-SNR in >20% of retained epochs were removed.

Power spectral density was estimated using a multi-taper method with discrete prolate spheroidal sequence tapers (L = 3). Mean power was extracted for six frequency bands: delta (1-4 Hz), theta (5-8 Hz), alpha (9-12 Hz), beta (13-30 Hz), low gamma (31-80 Hz), and high gamma (81-150 Hz). For P1 and P3, gamma bands were adjusted (31-55 Hz; 71-110 Hz) to avoid contamination from stimulation artifacts at 130 Hz and its subharmonic (65 Hz). Sham stimulation in P1 and P3 was delivered at the target stimulation frequency with minimal current (0.05 mA). In P2, sham trials were delivered through a second stimulator with the contacts left unconnected, so no current reached the brain, the recordings contained no 130-Hz or 65-Hz artifact, and the gamma bands were not adjusted.

Artifact-free gray-matter contacts were retained following the inclusion-exclusion criteria described in the QSA. Features (power of the six canonical frequency bands per channel) were log-transformed and z-score normalized within each recording day and used to train logistic regression classifiers to differentiate between desirable and undesirable symptom states.

#### Biomarker reliability and per-day decoding

Signals were re-referenced to same-region bipolar derivations, and spectral power was estimated for every bipolar contact through a multi-taper method for each two-second non-overlapping segment within one-minute intervals aligned to the per-minute VAS ratings, in 10-Hz bands covering the gamma range from 30 to 150 Hz. For each window the per-contact, per-frequency power values were log-transformed and normalized. Artifacts were rejected by robustly z-scoring each session’s log-power spectral features against their median and median absolute deviation, flagging any timepoint-contact-frequency entry whose absolute z-score exceeded 6, removing timepoints at which more than 10% of contacts were flagged and, among the remaining timepoints, contacts at which more than 2% of a day’s timepoints were flagged, and eliminating residual flagged entries with a linear program that selected the smallest set of timepoints and contacts forming a vertex cover of the bipartite contact-by-timepoint violation graph.

Symptom states were derived from the six VAS rating scales, similarly to the previous analysis. Within each patient and recording day, every dimension was normalized against a fitted normal cumulative distribution over the central 5 to 95 percent of its range, or by mean-centering dimensions with low variance. The normalized ratings were clustered with k-means into two clusters, similarly to the procedure described earlier. The clusters were labeled high-OCD and low-OCD and used as the binary decoding target.

A factorized logistic-regression classifier was fit separately for each patient. The classifier represents the contact-by-frequency weight matrix as the outer product of a contact-importance vector and a frequency-importance vector. For each window the two importance vectors were multiplied with the feature matrix to yield a single scalar, was passed through a sigmoid function to predict the label. The two vectors were optimized to maximize the log-likelihood of the observed labels to predict the data. Optimization used the AdamW optimizer with a learning rate of 0.1 and a weight decay of 0.1, run for 30 epochs over batches of 100 windows drawn by a random sampler.

Model performance was evaluated with iteratively stratified cross-validation, repeated over 10 random seeds. Folds were stratified jointly by the normalized VAS ratings, and windows from the same one-minute rating interval were kept in the same fold. For each seed, AUROC was computed separately for each recording day and then averaged across days. Reported per-day AUROC values are means across seeds, and error bars show the standard deviation across seeds.. For P1, whose provocation sessions were shorter, days were pooled into two groups (days 3, 4, 5, 7 and days 8, 10, 11) to reach the minimum number of ratings per class for the AUROC to be interpretable. Models were fit within patient by design.

To identify regional biomarkers, the model was bootstrapped 100 times on the pooled training, validation, and test data, and the learned importance scores were averaged within anatomical brain regions. For each region a bootstrap p value was computed as the smaller of the number of positive and the number of negative importance values across the 100 runs, divided by the number of runs. Regions whose Benjamini-Hochberg FDR-adjusted p value was at or below 0.05 were classified as biomarkers.

#### Acute stimulation testing

Electrode contact configurations for acute stimulation testing were selected based on proximity to established DBS targets, patient-specific tractography, and effective connectivity results. Following an initial safety and tolerability assessment, stimulation was delivered using a g.Estim PRO stimulator (g.tec medical engineering GmbH, Schiedlberg, Austria) across a range of frequencies (5-130 Hz), pulse widths (60-90μs), and amplitudes (1-6mA) for durations of 1-20 minutes. Stimulation was administered under multiple conditions, including double-blinded, sham-controlled trials and during OCD symptom provocation. Multiple contact configurations targeting a given region (e.g., cACC, ALIC, STN) were tested for practical reasons (e.g., high-impedance contacts). Stimulation trials within a region were combined for analysis, as were single-pulse responses for the effective-connectivity analysis.

#### Biomarker changes with stimulation

Because too few trials were available per target, stimulation of ALIC and NAc/VeP contacts was pooled as ventral capsule/ventral striatum (VC/VS) stimulation. Intracranial EEG recordings during acute stimulation were analyzed for changes in spectral power. For each trial, data from 10 seconds before stimulation onset and 13 seconds after stimulation offset were extracted. The first three seconds after stimulation offset were excluded due to hardware artifacts. Data were preprocessed as above and re-referenced using common-average referencing by least anticorrelation (CARLA^58,59^) and epoched to one-second long segments without overlap. Detrending was applied to these segments.

Analyses were restricted to frontal (OFC and FP) low-gamma contacts selected in more than 90% of symptom-state classifier runs (**Fig. 2a**). P3 had no such contacts, so its most frequently selected frontal low-gamma contacts were used instead (OFC, 41% of runs; FP, 28%); the FP contact did not survive artifact rejection. Epochs with stimulation artifacts were excluded using amplitude (see exclusion criteria in QSA). Power spectral density was computed for each frequency band, and contact-level power values were z-score normalized within stimulation session to facilitate comparisons across sessions. Normalization was performed by session to account for changes in impedance values throughout the day induced by stimulation.

#### Effective connectivity

Effective connectivity was assessed by sequentially stimulating all adjacent electrode contact pairs in bipolar configuration with 30 biphasic pulses (200 μs, 6 mA, inter-pulse interval 3-7 s) while recording from all other contacts. Signals were high-pass filtered at 0.5 Hz and epoched from 0.5 s pre-stimulation pulse to 2 s post-stimulation pulse. Weak connections were thresholded as described in the QSA (see Statistical robustness analyses). Recordings were re-referenced on a per-headbox basis using common average referencing by least anticorrelation (CARLA ^58,59^).

For each stimulation-recording pair, the evoked responses were averaged across trials. The response amplitude was computed as the root-mean-square (RMS) of the z-scored signal in 10-500 ms post-stimulus window, normalized to the 500-10 ms pre-stimulus baseline. Subnetwork EC values were computed as the product of individual connections. Associations with acute clinical changes were assessed as described in the QSA.

#### Patient-specific tractography

High-resolution MRI sequences were collected for each patient: T1-weighted Magnetization-Prepared Rapid Gradient-Echo (MPRAGE) with and without gadolinium contrast, T2-weighted, and Fast Gray Matter Acquisition T1 Inversion Recovery (FGATIR) sequences. Multi-shell diffusion-weighted MRI (b=1500, 3000) was acquired at 1.5mm isotropic voxels and 64 diffusion directions, along with a reversed phase-encoding b0 (blip-down) image. DWI was pre-processed using QSIPrep^60^, followed by estimation of voxel-wise fiber orientation distributions using FSL’s BEDPOSTX^61^ and co-registered to the structural T1-weighted imaging using boundary-based registration. Atlas-based anatomical segmentation was performed following a two-step linear and non-linear co-registration using Advanced Normalization Tools^62^ framework with affine and asymmetric normalization. Atlas-defined region-of-interests (ROIs) were transformed into native diffusion space and inspected visually to ensure accurate anatomical localization. Probabilistic tractography was performed using FMRIB Software Library’s Probtrackx2 with 5000 samples per seed voxel from each seed region to target ROIs ^63^. For each seed voxel, 5000 streamlines were initiated and propagated stepwise according to the local posterior probability distributions of fiber orientation estimated by BEDPOSTX. Streamlines were terminated upon reaching a curvature threshold or exiting brain mask. Connectivity distributions were generated by accumulating the number of samples reaching each target ROI. DTI-based connectivity was quantified using normalized waytotal (i.e., waytotal divided by the total number of generated streamlines). Normalized waytotals for subnetworks containing more than two nodes were computed by multiplication, as above. See QSA below for a description of correlations with acute clinical changes.

#### Pathway-activation modeling

Pathway-activation modeling relies on two key components. First the structural connectome model and second the biophysical stimulation model. These two components are integrated to simulate the cortical connectivity associated with subcortical stimulation^64^.

Our structural connectome model was defined by Mehta et al.^42^. The Mehta internal capsule connectome was created using the i80 ex-vivo human brain of the Large-scale Imaging of Neural Circuits (LINC) dataset^65^. Their pathway reconstruction methods consisted of blending traditional tractography with cortical innervation stitching to uniformly sample internal capsule tract connectivity with the entire frontal cortex. The Mehta pathways are a stylized connectome of 6000 total tracts, which were individually curated to match known histological axonal trajectories through the internal capsule^66,67^. They include representation from eight prefrontal cortical territories associated with the anterior limb of the internal capsule (ALIC) and five motor territories associated with the posterior limb. Twenty percent of the cortical tracts exhibit a collateral branch into the STN, representing the hyperdirect pathway^24,68^. The tracts uniformly sample every cortical territory associated with the frontal cortex and are defined within the coordinate system of the CIT168 human MRI brain atlas^69^. We then used symmetric normalization (SyN) in ANTs^70^ to co-register the CIT168 brain atlas to each patient MRI, thereby placing the Mehta pathways into patient-space.

Accurately predicting the neural response to the voltage distribution generated by subcortical stimulation is an especially difficult challenge^71,72^. For generalizability, we elected to employ the simplest possible concept for simulating an empirical activation volume, which is known as the current-distance relationship^73^ (I_th_(pw, d_axon_) = k * r^X^). We generated our best fits to this analytical equation using the massive library of pathway-activation results from Frankemolle-Gilbert et al.^74^. The fitted model provided an estimated activation volume for each experimentally tested stimulation parameter setting. The overlap between the stimulation volume and the Mehta pathways (in patient-space) provided a collection of cortical tracts for analysis for each tested setting. The fraction of ALIC tracts terminating in the OFC was taken as the pathway-activation measure used in **Fig. 6a**. Correlations with acute clinical changes are described in the QSA below.

#### DBS implantation and therapy

Following sEEG explantation, patients underwent implantation of a four-lead, rechargeable DBS system (Vercise Genus 32, Boston Scientific, Marlborough, MA). Leads were placed in personalized targets informed by sEEG-derived subnetwork analyses. Lead placement was confirmed with intra- and post-operative CT scans. Patients underwent biweekly DBS optimization for up to 52 weeks (60 weeks for P1). The last common timepoint across patients was 32 weeks. Hemispheres were activated sequentially to assess efficacy and tolerability. Clinical response was measured using the Y-BOCS-II.

### QUANTIFICATION AND STATISTICAL ANALYSIS

#### General

Unless otherwise specified, statistical details for each analysis (test used, exact *n* and what it represents, definition of center and dispersion, and significance thresholds) are reported in the corresponding figure legends and below. Given the cohort size (N=3), analyses relied on non-parametric and permutation-based inference rather than parametric tests. No formal normality testing was performed. Two-sided tests were used throughout unless stated. Significance markers denote *** p < 0.001, ** p < 0.01, * p < 0.05. Spectral preprocessing and feature extraction were performed in MATLAB, using Fieldtrip ^56^ (v20240620) and custom code, or Python (v3.11.11); statistical analyses were performed in Python (v3.11.11; scikit-learn ^75^ v1.5.2, SciPy, statsmodels).

#### Sample size, randomization, blinding

No formal sample-size estimation or power analysis was performed. N was constrained by the available SEEG cohort. Acute stimulation testing included double-blinded, sham-controlled trials. Sham stimulation in P1 and P3 was delivered at the target stimulation frequency with minimal current (0.05 mA); P2 received no stimulation during sham trials, but the stimulator was activated at the target stimulation frequency to produce the sound of stimulation. High-versus low-symptom class labels were derived per trial by k-means clustering of self-reported visual-analog symptom ratings (scikit-learn), with cluster centroids reviewed clinically and classified as desirable or undesirable. For P3, whose ratings were highly correlated across scales and sometimes missing, labels were instead defined by thresholding distress ratings. Number of clusters was confirmed using silhouette analysis. Labels were used for all symptom-state decoding and power-difference analyses.

#### Data inclusion and exclusion

For spectral biomarker analyses (**Fig. 2**), artifact-contaminated channels and epochs were identified semi-automatically. Low-SNR contacts were detected with the NoiseTools library; epochs containing >20% low-SNR contacts were excluded, and contacts marked low-SNR in >20% of retained epochs were removed. Only artifact-free gray-matter contacts were retained. Per-feature outliers were detected by the interquartile-range method (IQR multiplier k = 2) and imputed by k-nearest neighbors (k = 5). For acute-stimulation power analyses, epochs were excluded by amplitude threshold (P1: >100 μV; P2, P3: >115 μV), and contacts exceeding a contamination fraction (P1: >20%; P2, P3: >25%) were removed; when only a single trial was available in either comparison group, no statistical test was performed. For effective connectivity (**Fig. 4**), non-significant connections and connections <15 mm apart were set to zero prior to analysis.

#### Effective connectivity

For each stimulation–recording pair, evoked-response amplitude was computed as the RMS of the z-scored signal in a 10-500 ms post-stimulus window, normalized to a 500-10 ms pre-stimulus baseline and averaged across trials (**Fig. 4**). Significance of each connection was assessed by Wilcoxon signed-rank test with Benjamini–Hochberg correction (α = 0.01). Resulting associations with acute clinical change were assessed by weighted Pearson correlation as above (shown in **Fig. 4**).

#### Connectivity-response associations and statistical robustness

Associations between connectivity measures and acute clinical response were quantified with weighted Pearson correlations, using the number of stimulation trials per observation as weights this same weighted-correlation procedure was applied to effective connectivity, DTI-based normalized waytotals, and the activated ALIC-to-OFC streamline measure ^76^. Subnetwork values (>2 nodes) were computed as the product of constituent connections. Acute associations were evaluated across the six implanted hemispheres (two per patient). Significance was confirmed non-parametrically by permutation testing: clinical-response labels were randomly shuffled across observations 720 times (all possible permutations), the statistic recomputed for each permutation, and the empirical P value defined as the proportion of permutations with a statistic at least as extreme as observed. Robustness of the acute hemisphere-level correlations (connectivity and tract activation) was assessed by refitting each correlation after leaving out each hemisphere and, separately, each patient, and reporting the mean and range of R² across refits. Leave-one-out influence diagnostics confirmed that no single observation drove the reported correlations.

#### Symptom-state classification

High-versus low-symptom states were discriminated per patient with a binary L2-regularized logistic-regression classifier (scikit-learn), evaluated over 200 repetitions of grouped, stratified two-fold cross-validation^77,78^; within each repetition the two complementary folds were averaged to yield 200 held-out AUC estimates per patient, summarized as the mean AUC (**Fig. 2a**). Feature selection (univariate ANOVA, top 20 features) was performed within each training fold only. Significance was assessed against a matched null of 200 label permutations, each shuffling class labels before fitting and averaging over its two folds ^79^; the permutation p-value was 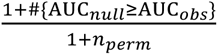, with *n_perm_* = 200. Here *n* = 200 repetition-level AUCs per patient model; the underlying units are within-patient, time-resolved power samples (segments per patient: P1, 105; P2, 413; P3, 235), and models were fit within patient and not pooled (N = 3 patients).

#### State-dependent power differences

Power differences between high- and low-symptom states were tested per feature with a two-sided permutation test on the difference in mean z-scored power, using 10,000 label shuffles and the p-value computed as 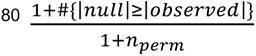 – results shown in **Fig. 2a**. Within each patient panel, raw p-values across the displayed features were corrected for multiple comparisons by the Benjamini–Hochberg FDR procedure (α = 0.05), and significance markers reflect the corrected values: *** p < 0.001, ** p < 0.01, * p < 0.05, else n.s. For every feature, n is the number of z-scored power samples (day–block–trial epochs) in each state and is printed beneath each box; boxes show the median (center) and interquartile range with whiskers at 1.5 × IQR, and all observations are overlaid. Tests were within patient (N = 3 patients; SciPy, statsmodels).

#### Biomarker robustness and per-day decoding

The high symptom state was decoded with a factorized logistic-regression model applied to multitaper spectral power in 10-Hz bands (30–150 Hz) over 2-s windows of bipolar-referenced, artifact-screened sEEG, with per-trial high/low labels derived by k-means as above. Robustness was evaluated with stratified day-wise splits, with performance summarized as the average per-day AUROC. Details concerning data preprocessing and cleaning are described in STAR Methods.

#### Acute stimulation responses

For each stimulation site, acute changes in each VAS scale were compared with the patient’s sham trials (a single sham condition per patient; see above) using one-sided Wilcoxon rank-sum tests, which test specifically for a decrease in symptom ratings relative to sham. Given the exploratory, within-patient design, p-values were not corrected for multiple comparisons across sites and scales. Transition probabilities were defined on the multidimensional symptom states (k-means clusters over all six VAS scales for P1 and P2, thresholding of distress for P3; see Sample size, randomization, blinding): for each site, the proportion of trials that moved the patient from the high-to the low-severity state, and separately from the low-to the high-severity state (**Fig. 3b**). Transition probabilities were compared with sham descriptively.

#### Biomarker changes with stimulation

Power changes were computed per trial as post-minus-pre stimulation power (contact-level values z-scored within stimulation session to control for impedance drift) and compared between trials associated with symptom improvement versus worsening by two-sided permutation testing (1,000 permutations, α = 0.05). No test was performed when only one trial was available in either group (P2, OFC).

## Data Availability

De-identified data and analysis code are available on GitHub

https://github.com/Halpern-Scangos-Lab/Personalized-Network-Biomarkers-of-DBS-Response-in-OCD

## Acknowledgments

This work was supported by the Rodan Family Foundation, the AE Foundation, and the New Venture Fund (Foundation for OCD Research Seed Grant). R.L.S. was supported by grants R25MH119043, T32NS091008, T32DA028874. L.Q. is supported by the Brain & Behavior Research Foundation as the Ellen Schapiro & Gerald Axelbaum Investigator. C.C.M, A.M.N., and H.C. were supported by NIH R35 NS143008. The authors would like to thank the study subjects for their participation and commitment to this novel, exploratory application.

