## Supplemental Figures 1-5 and Supplemental Table 1 for "Patient-specific subnetwork connectivity predicts deep brain stimulation response in obsessive-compulsive disorder"

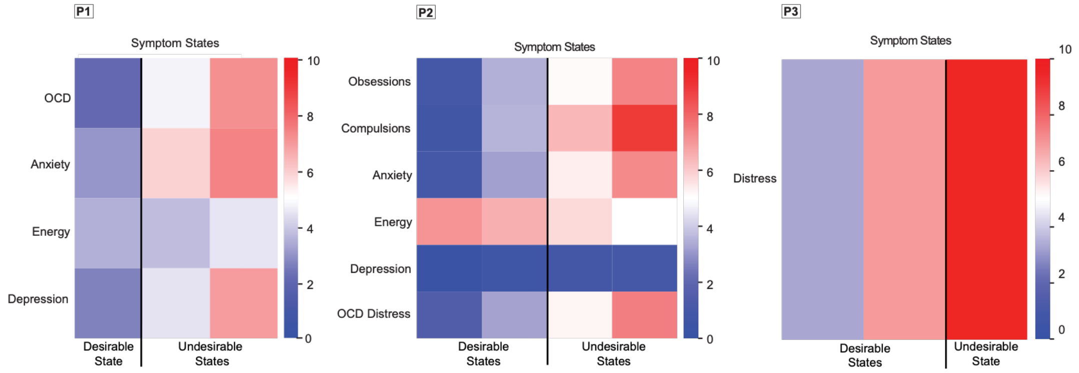


**Figure S1. Patient- and target-specific changes in symptom state.**
Heatmaps showing symptom states for each patient, classified as desirable or undesirable. States were defined using k-means clustering of all symptom ratings for each patient (total ratings: P1: 597; P2: 755; P3: 724). For P1, the obsession and compulsion VAS scales were averaged to form a combined OCD rating because the values were two scales were highly correlated. For P3, symptom states were defined using distress ratings, the only symptom measure available for many trials.

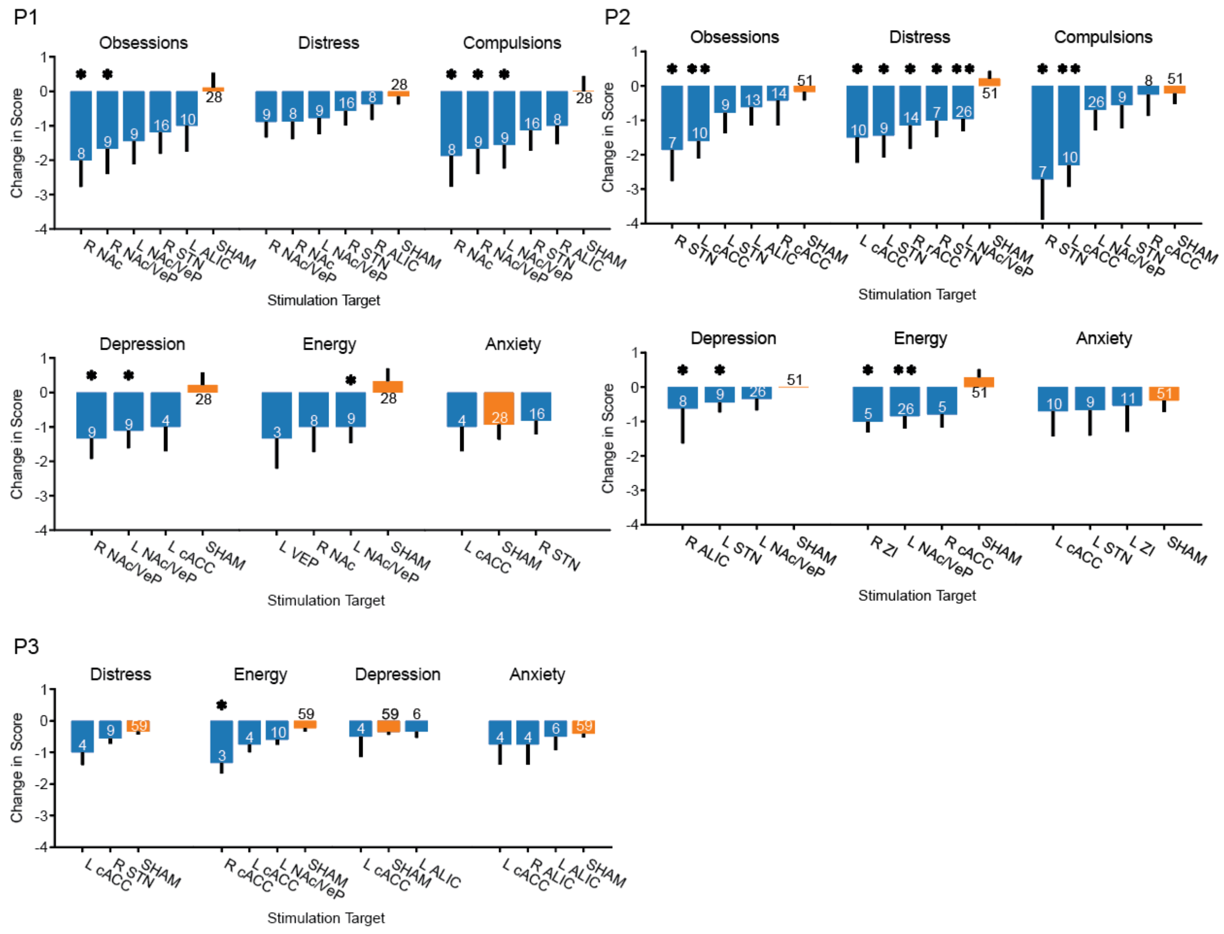


**Figure S2. Differential clinical response to stimulation by target and patient**

Per-patient bar plots showing stimulation sites associated with the greatest mean change in symptom ratings for each symptom scale, alongside sham stimulation. For each scale, stimulation sites with mean symptom reduction greater than sham are shown in decreasing order (blue), with the mean change during sham stimulation shown for comparison (orange). Number on bars indicate the number of stimulation trials per target, and one-sided error bars represent standard error of the mean. Symptom reductions significantly greater than sham are indicated by * and ** ($p<0.05$ and $p<0.01$, respectively; one-sided Wilcoxon rank-sum test).

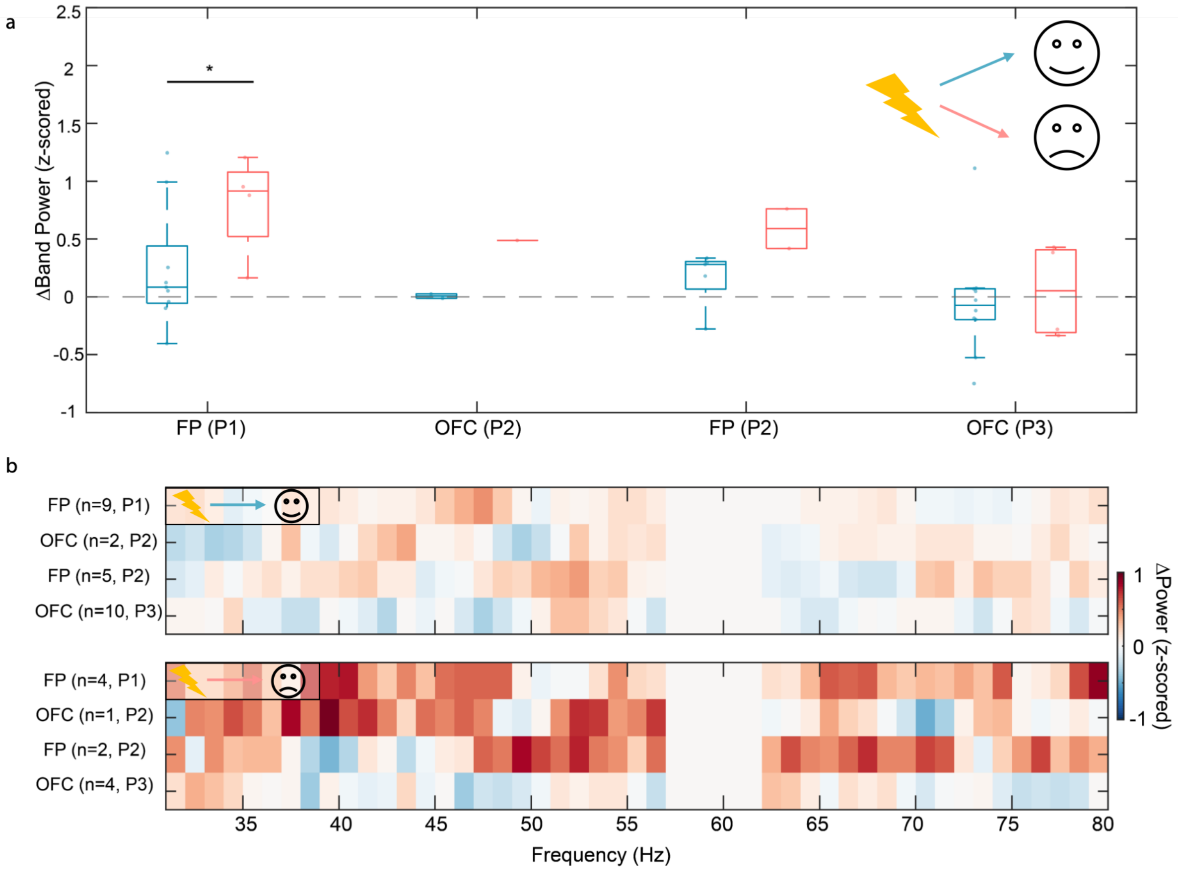


**Figure S3. Stimulation-induced frontal gamma power changes associated with induced symptom changes** a) Box and whisker plot of gamma band power changes per patient and frontal region (OFC - orbitofrontal cortex, FP – frontal pole) in response to VC/VS stimulation that induced improved symptom state (blue) and worsening symptom state (red). b) Heatmaps of VC/VS stimulation-induced changes in gamma power in frontal regions for each patient and in response to stimulation that improved (top) and worsened (bottom) symptom state.

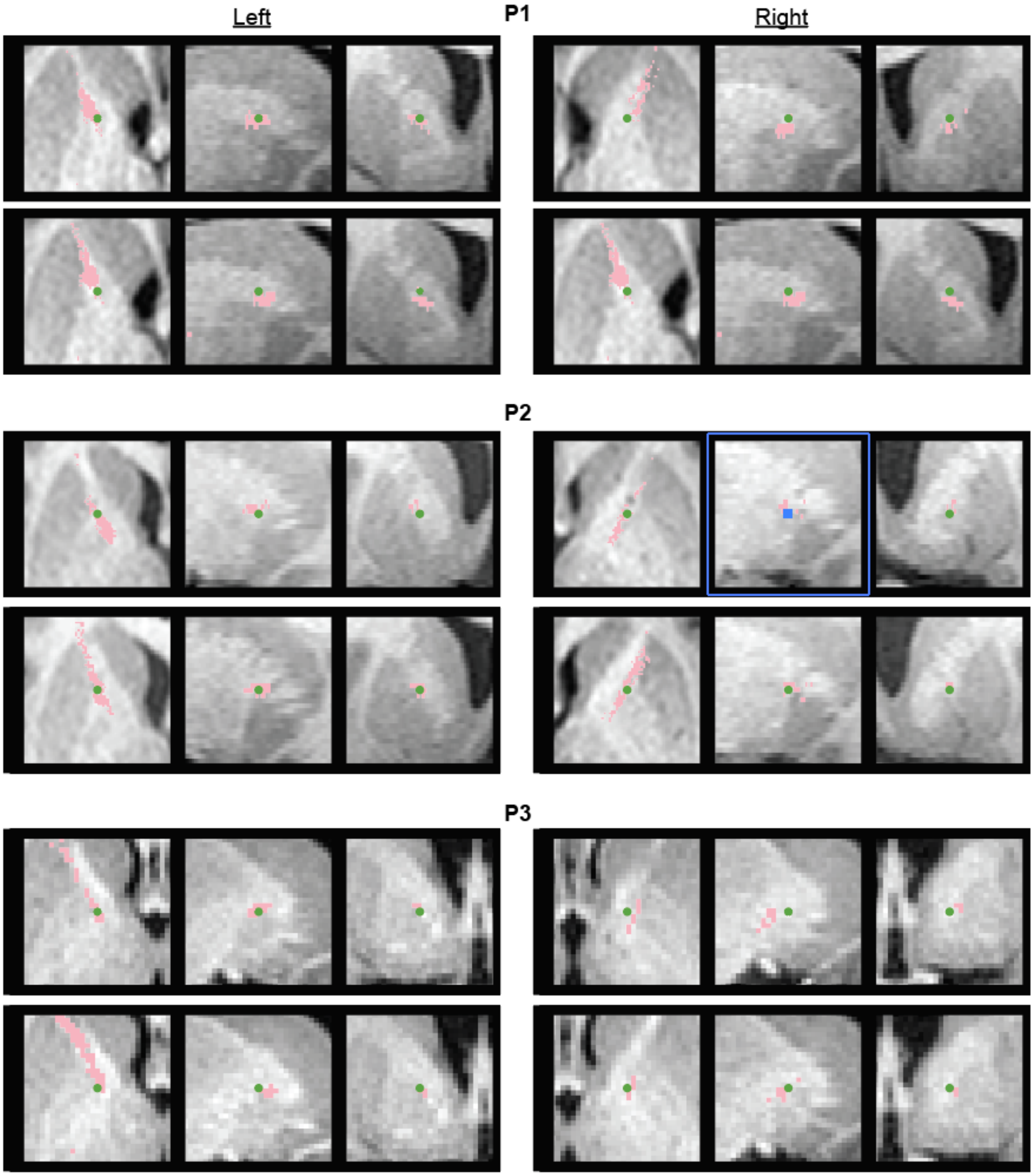


**Figure S4. ALIC contact localization.**
Axial, sagittal, and coronal CT sections from each hemisphere showing electrode contact locations used for effective connectivity analyses (green) within the anterior limb of the internal capsule (ALIC), in proximity to streamlines correspond to the “OCD response tract” (pink) for each patient.

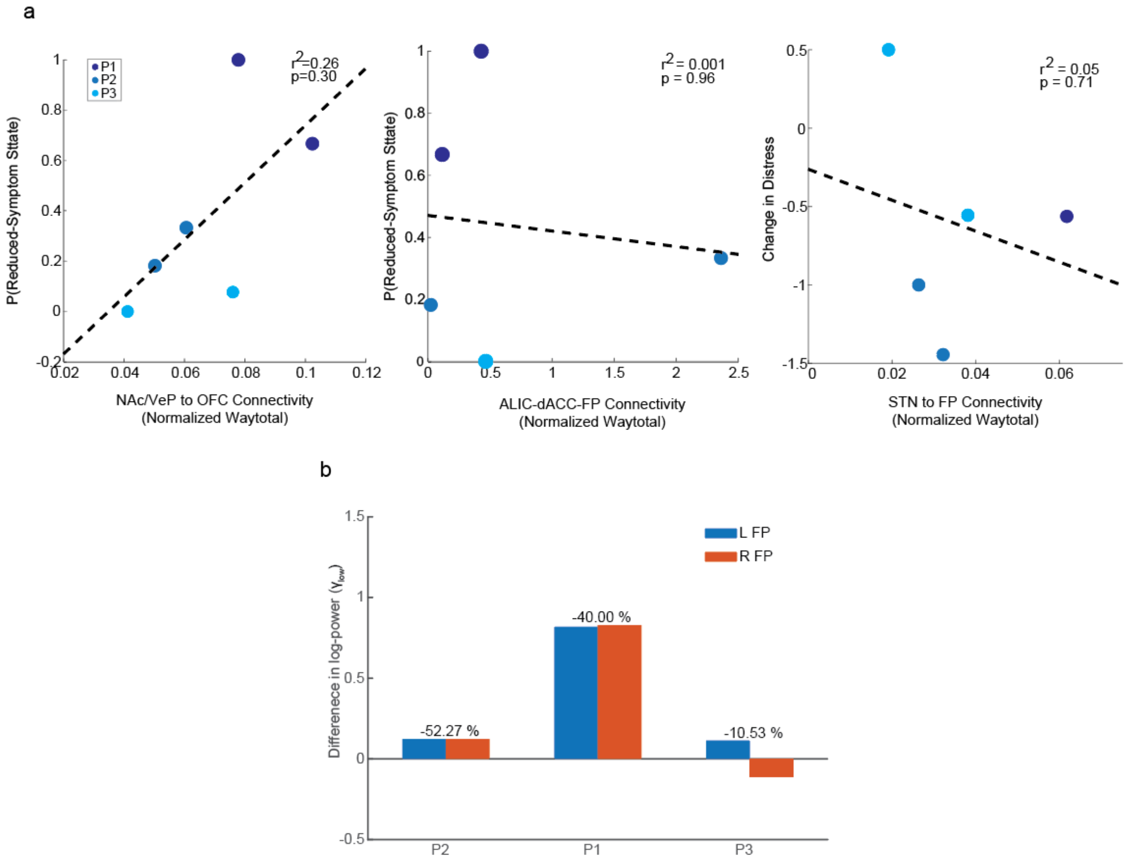


**Figure S5. Structural connectivity and spectral state biomarkers do not predict stimulation-related clinical improvement.**

**a,** Scatter plots showing no significant association between subnetwork connectivity measures by diffusion tensor imaging (DTI)-derived normalized waytotal and acute OCD symptom improvement. Dashed lines indicate the weighted linear fit. **b,** Frontal pole gamma power derived from the spectral activity biomarker shows no clear association with the percent change in Y-BOCS-II during chronic DBS (percent change indicated above bars).

**Supplemental Tables:**

| Patient | Age Range | Sex | Primary OCD Symptom | Baseline Y-BOCS/ Y-BOCS-II | OCD Medications at Enrollment |
|---|---|---|---|---|---|
| P1 | 35-40 | Male | Chronic doubting | 30/40 | Paroxetine 20mg<br>Quetiapine 25mg |
| P2 | 25-30 | Female | Contamination | 34/44 | Sertraline 100mg<br>Trazodone 100mg<br>Clonazepam 0.5mg |
| P3 | 60-65 | Female | Contamination | 29/38 | Levomilnacipran 120mg<br>Aripiprazole 2mg<br>Lisdexamfetamine 70mg |

**Table S1.** Baseline patient demographics, OCD characteristics, and treatments
